# How effective are UK-based support interventions and services targeted at adults who have experienced domestic and sexual violence and abuse at improving their safety and wellbeing? A systematic review protocol

**DOI:** 10.1101/2023.07.14.23292666

**Authors:** Sophie Carlisle, Annie Bunce, Matthew Prina, Sally McManus, Estela Barbosa, Gene Feder, Natalia Lewis

**Affiliations:** Department of Health Service and Population Research, Institute of Psychiatry, Psychology and Neuroscience, King’s College London, De Crespigny Park, London, UK; Violence and Society Centre, City, University of London, London, UK; Population Health Sciences Institute, Faculty of Medical Sciences, Newcastle University, Newcastle, NE4 5PL, UK; National Centre for Social Research, London, UK; Centre for Academic Primary Care, Bristol Medical School, University of Bristol, Bristol BS8 2PS, UK

## Abstract

**Background:** Domestic and sexual violence and abuse (DSVA) is prevalent in the UK, with wide-ranging impacts both on individuals and society. However, to date, there has been no systematic synthesis of the evidence for the effectiveness of UK-based support interventions and services for victim-survivors of DSVA. This review will aim to systematically collate, synthesise and quality assess the evidence regarding the effectiveness of UK support interventions and services targeted at those who have experienced DSVA. The review will use findings of a preliminary scoping review, as well as input from stakeholders representing domestic and sexual violence third sector organisations to identify and prioritise the most relevant outcomes to focus on.

**Methods:** We will undertake a systematic search for peer-reviewed literature in MEDLINE, EMBASE, PsycINFO, Social Policy and Practice, Applied Social Sciences Index and Abstracts (ASSIA), International Bibliography of the Social Sciences (IBSS), Sociological abstracts and SSCI. Grey literature will be identified by searching grey literature databases, circulating a call for evidence to local and national DSVA charities and organisations, and targeted website searching. Two reviewers will independently perform study selection and quality appraisal, with data extraction undertaken by one reviewer and checked for accuracy by a second reviewer. Narrative synthesis will be conducted, with meta-analysis if possible.

**Discussion:** Existing individual studies and evaluations have reported positive impacts of support interventions and services for those who have experienced DSVA. Thus, it is expected that this review and synthesis will provide robust and conclusive evidence of these effects. It will also allow comparisons to be made between different types of support interventions and services, to inform policy makers and funders regarding the most effective ways of reducing domestic and sexual violence and abuse and its impacts.

## Introduction

Domestic and sexual violence and abuse (DSVA; see Appendix 1 for glossary definition) is a prevalent international public health issue, disproportionally affecting women and girls. Domestic abuse has been estimated to affect 1 in 3 women and 1 in 7 men in England and Wales in their lifetimes [1], and nearly 1 in 4 women and 1 in 20 men have been victim to some form of sexual assault since the age of 16 [2]. Whilst already high, these are likely to be underestimates, with social stigma [3] and the context the survey is delivered in [4] influencing estimates. Whilst data on perpetrators are not routinely collected in the UK currently, several cross-sectional studies have investigated this, with 20% of men reporting perpetration of intimate partner violence and abuse by the age of 21 years in one UK study [5], and another multi-country study reporting prevalences of intimate partner violence perpetration by men aged 18-49 years that ranged from 25.4% in rural Indonesia to 80% in Papua New Guinea [6]. The adverse impacts of DSVA are plentiful and varied both for those experiencing DSVA, and for the wider society. DSVA affects physical health [7–13], mental health [4, 14–16], financial stability, employment, education [17], homelessness [18], relationships, and substance dependence [19]. DSVA also puts significant demand on services, such as the criminal justice system [20], health and social services [21] and police [22]. Collectively, the total cost of domestic abuse for society considering all of these consequences has been estimated at approximately £66 billion over a one-year period [23].

Support services and interventions play an essential role in meeting the diverse needs of those affected by DSVA. These may include refuges, advocacy, referral, outreach, counselling, financial or legal advice, and helplines, to name a few. In the UK, such services and interventions are often provided by third sector organisations, however, they may also be located in both the public and private sector. Evidence from previous systematic reviews of the global evidence, focusing on one or more of these interventions and services suggests that they are effective in providing support. For instance, housing interventions have been found to improve mental health and perceived safety and reduce stress [24], and economic interventions have been shown to reduce levels of domestic violence and increases in empowerment [25]. Similarly, a review of advocacy interventions showed improvements quality of life and depression [26], whilst a review of psychological therapies showed benefits in terms of depression and anxiety [27]. Reviews of perpetrator programmes (i.e., interventions targeted at people who use violence) have also shown some benefits in terms of reduction in the perpetration and experience of abusive behaviours [28, 29]. Thus, there is some existing evidence that support interventions and services lead to improvements in a range of outcomes. Whilst the exact mechanisms through which these changes occur are unclear, it may be related to the provision of resources, whether that be practical (i.e., housing, financial support), psychological (i.e., increased coping and resilience, space to process trauma) or informational (i.e., information about other services, available options, and next steps). Together, these may improve mental health, wellbeing, and feelings of empowerment, enabling those experiencing DSVA to be in a better position to achieve their own goals [30, 31] and live a life free from abuse.

However, whilst the effectiveness of such interventions and services have been evaluated separately, to date no review has assessed the evidence for support interventions and services in the UK as a whole. One issue facing such a synthesis is the wide-ranging outcomes used to assess effectiveness. A core outcome set across services and intervention types has not been agreed, and currently many different outcomes and outcome measures are being used. Indeed, we recently undertook a scoping review of all effectiveness outcomes used in studies and evaluations of support interventions and services for people with experience of DSVA and found a total of 426 outcomes across 80 studies, with only 46.9% used more than once [32]. The outcome measure used most frequently by the included studies was the cessation of abuse, a proxy measure of safety, most commonly assessed using the Severity of Abuse Grid [33]. Presence of abuse, severity of abuse (also proxy measures of safety) and self-esteem measured by the Rosenberg Self-Esteem Scale were also identified as commonly reported outcomes.

## Objectives

The aim of this review is to synthesise the evidence of the effectiveness of support services and interventions for people experiencing DSVA on safety and wellbeing, specifically focusing on outcomes identified by in the scoping review and by our stakeholder advisory group. In doing so, we hope to identify which interventions and services are most likely to benefit those who have experienced DSVA in these areas. This may be particularly helpful for third sector organisations with limited funding to maximise their benefit, and provide evidence to support future funding applications. The identification of efficacious interventions and services may also encourage greater investment from external funders and inform future cost-benefit analyses to provide further justification. Finally, this review will identify gaps in the evidence where effectiveness studies or evaluations are needed, and if/where higher quality studies of the effectiveness of specific types of services or interventions are needed.

## Review question

1. How effective are UK-based support interventions and services (targeted at adults of any gender who have experienced DSVA) at improving safety and wellbeing?

## Methods and analysis

This protocol follows the Preferred Reporting Items for Systematic Review and Meta-Analysis Protocols (PRISMA) checklist (Appendix 2) [34]. The systematic review will follow PRISMA guidelines [35]. The protocol has been registered on Prospero: CRD42022339739.

## Eligibility criteria

We will select intervention studies according to the following criteria (Table 1).

**Table 1.**
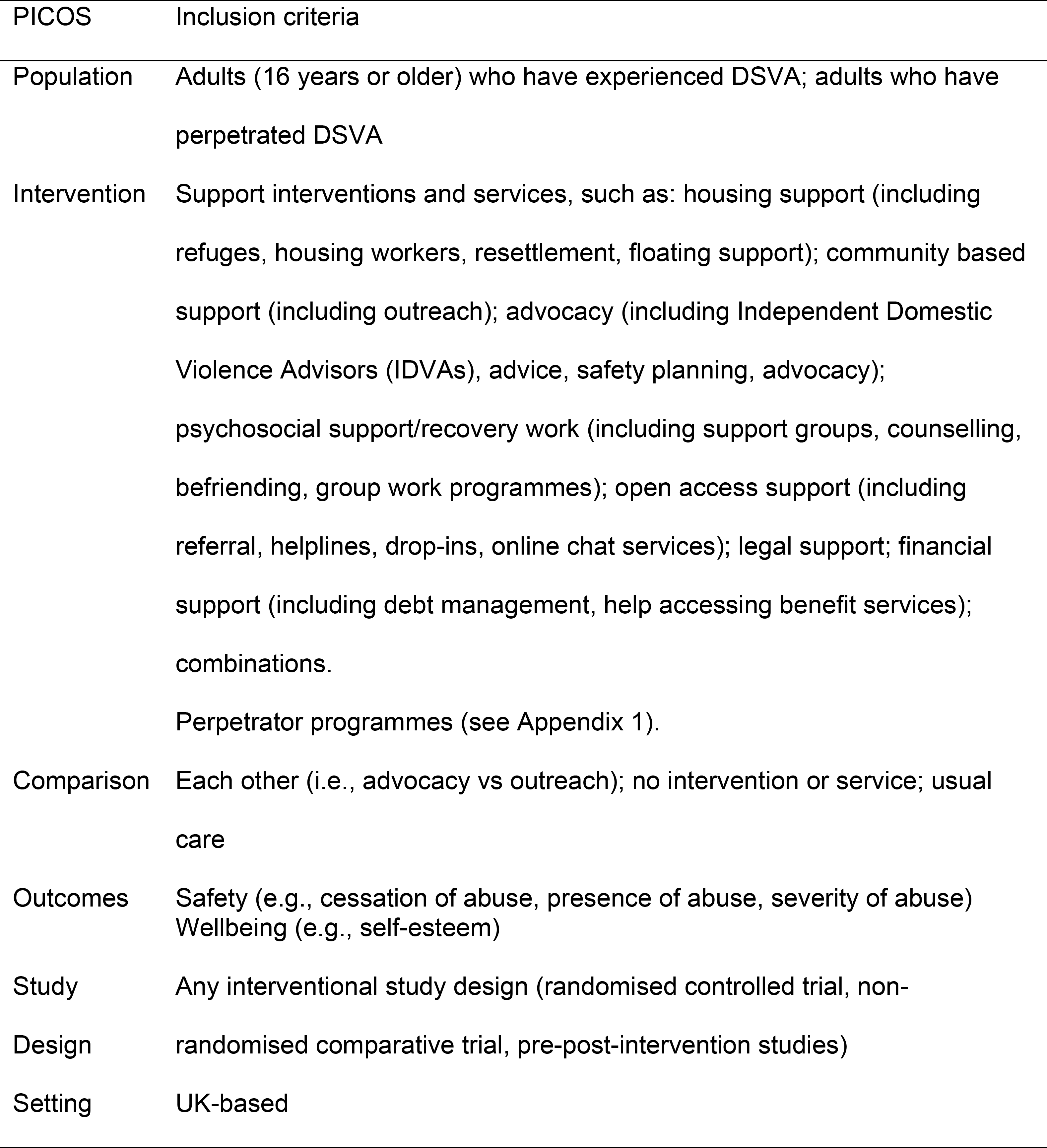
Inclusion criteria.

### Population

Adults of any gender who have experienced DSVA, or who have perpetrated DSVA. Adults are defined as those aged 16 years and older, in line with the definition used in the National Institute for Health and Care Excellence quality standard [36]. It should be noted that in this review we use the term ‘people who have experienced DSVA’, however due to different terminology preferences between organisations within the third sector, this may also be used interchangeably to mean victims of DSVA, survivors of DSVA, and victim-survivors. We are not placing a limit on time since the experience of violence, so long as participants accessed an intervention or service as an adult.

If the population of a study is mixed (e.g., studies that include a mix of individuals who have experienced DSVA and a form of violence other than DSVA), the study will only be included if the proportion of participants meeting the inclusion criteria is more than 50%, or if outcomes are reported separately for the populations.

In this review, domestic violence and abuse (DVA) has been defined according to both the UK cross-governmental (2013) [37] and the Domestic Abuse Act 2021 [38] definitions. Definitions of sexual violence and abuse (SVA) that will be covered by this review include the definitions given by the Istanbul Convention (Article 36) [39], the World Health Organisation [40] and the Rome Statute of the International Criminal Court’s (ICC) Elements of Crimes (2013) [41]. It is important to be aware of overlaps that exist between definitions. For instance, rape may be considered both domestic and sexual violence, or may be sexual violence only, depending on the relationship between the person using and the person experiencing violence. Similarly, intimate partner violence (IPV) is a form of domestic violence that may or may not include forms of sexual violence. Given the gendered nature of both domestic and sexual violence, some organisations use the term ‘violence against women and girls’ to encompass all forms of violence disproportionately experienced by women and girls, including domestic and sexual violence, as well as other forms of violence that may not be considered domestic or sexual. To illustrate the distinctions and overlaps between these definitions, we have developed a flowchart using examples of forms of violence and how they might be categorised (Figure 1).

**Figure 1.**
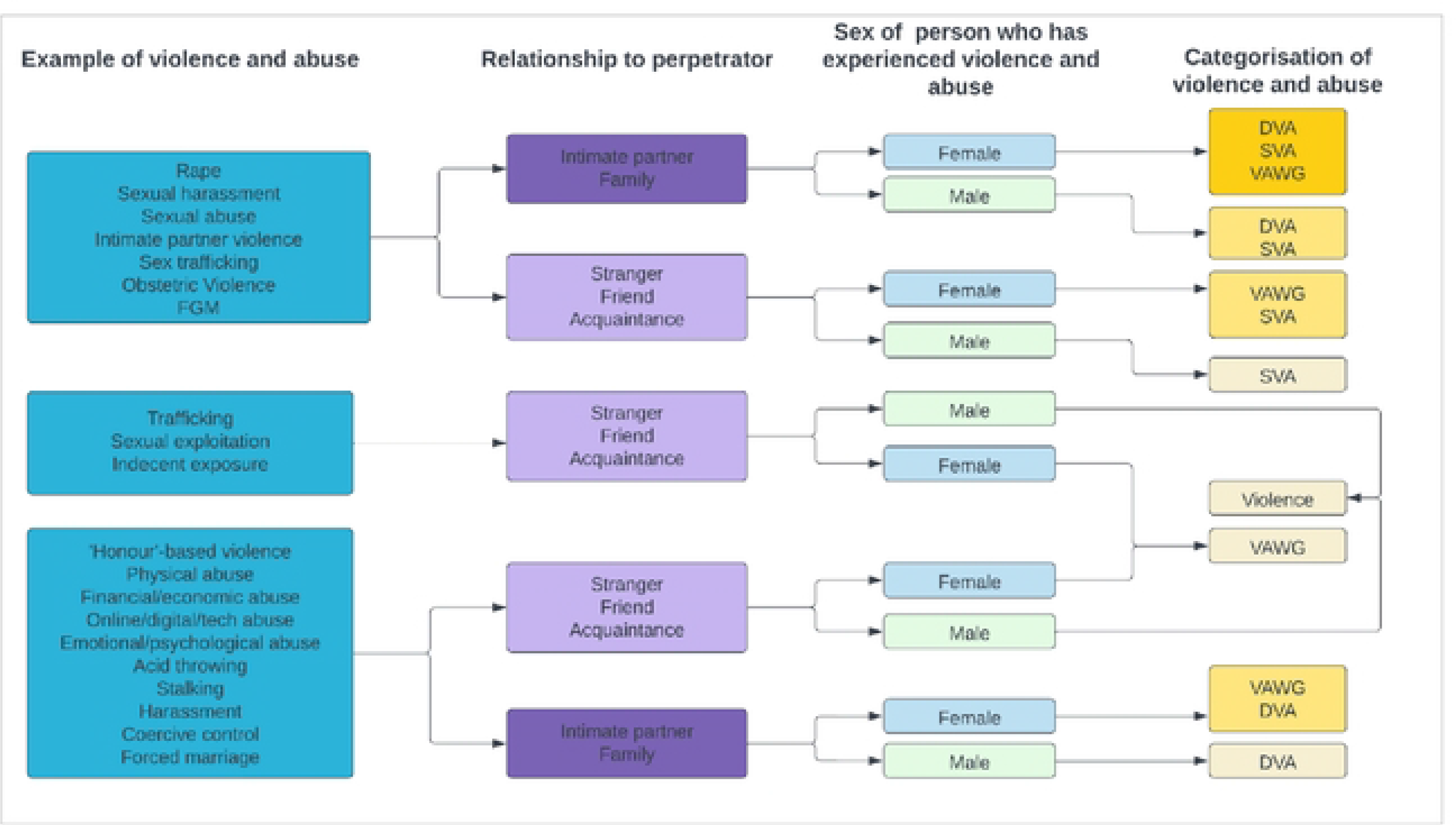
Flow chart to illustrate the relationship between forms of violence, the relationship between person who has experienced violence and perpetrator, the sex of the person who has experienced violence, and the categorisation of the form of violence. Note that these are examples, and this is not an exhaustive list. DVA: domestic violence and abuse; FGM: female genital mutilation; SVA: sexual violence and abuse; VAWG: Violence against women and girls.

### Intervention

Any secondary or tertiary prevention support interventions or service will be included, as these are aimed at those who have already experienced violence [42]. We will include multi-component interventions and services, and individual or group-based interventions and services, which may be of any duration. Entry to the intervention or service should be determined by the experience of DSVA (either as someone who has experienced or perpetrated DSVA). The types of support interventions and services eligible for inclusion include:

- Housing-related (refuges, housing workers, resettlement, floating support)
- Advocacy (advice, advocacy, safety planning, Independent Domestic Violence Advisers/Advocates (IDVAs), Independent Sexual Violence Advisers/Advocates (ISVAs))
- Community-based support (outreach)
- Psychological support/recovery work (support groups, counselling, befriending/peer support, group work programmes)
- Open access support (helplines, drop-in, referrals, online chat services)
- Legal support
- Financial support (debt management, help accessing benefit services)
- Multi-agency risk assessment conferences (MARACs)
- Police-based DSVA services

Additionally, perpetrator programmes of any nature will be included (see Appendix 1 for glossary definition). The inclusion of perpetrator programmes is based on several factors. Firstly, including them will allow the measurement of DSVA, including the cessation of abuse, without placing the burden on people who have experienced DSVA to change their perpetrators’ behaviour. This ties into the UK government’s new perpetrator strategy [43], which intends to place the onus of responses to DSVA on perpetrators changing their behaviour, alongside the recovery of those who have experienced DSVA. Finally, many perpetrator programmes offer associated support to (ex)partners or referral to appropriate support, and measure outcomes relating to their progress as well as that of the perpetrators.

Primary prevention interventions and services such as awareness raising and education will be excluded, as participants are not necessarily those who have experienced violence. Additionally, interventions or services that are not primarily aimed at DSVA will be excluded. Finally, the provision of online information as an intervention or service where there is no further contact with a named worker will not be included, because this does not constitute an active intervention, and may not be accessed only by those who have experienced or used DSVA.

### Comparator

The comparison intervention will be another included intervention or service, and usual care (including no support service or intervention or waitlist controls). In the case of pre-post study designs, studies without a comparison intervention will be included, as the comparison will be between changes from pre-intervention to post-intervention.

### Outcomes

We will include studies that report data on safety and / or wellbeing. The specific outcomes and outcome measures will be determined through two processes. Firstly, we will use the results of the scoping review that we conducted which mapped and charted all outcomes measuring effectiveness of support interventions and services, and identify the most commonly reported outcome measures. Secondly, we will consult with an advisory group (for details see Public and Patient Involvement) to determine which of the commonly reported outcomes identified by the scoping review are most relevant, useful and meaningful. This approach will ensure that the outcomes of the review are synthesisable, whilst at the same time ensuring that the review produces meaningful and useful information for third-sector services and researchers working in this sphere.

### Types of studies

Any type of interventional study, including randomised controlled trials (RCTs), non-randomised comparative trials, pre-post intervention studies and service evaluations will be included. Observational study designs, such as case-control, cross-sectional and case studies will be excluded, as will qualitative studies and theoretical studies.

### Setting

Any UK setting will be included (regardless of the immigration status or nationality of the service user or the location where the DSVA was experienced).

### Language

Given the focus of the review on UK-based specialist support services and interventions, only English language reports will be included.

## Search Strategy

Studies will be retrieved using eight databases: MEDLINE, EMBASE, PsycINFO, Social Policy and Practice, ASSIA, IBSS, Sociological abstracts and SSCI. Studies published before 01 January 1982 will not be included. The date limit is based on the year that the Crime Survey for England and Wales was first introduced. This is because it is hoped that the results of this review will be integrated with data from this survey as part of a wider programme of work. Searches will include terms relating to violence (e.g., “domestic violence”, “partner”, “sexual violence” etc.), support services and interventions (e.g., “specialist service”, “support”, “outreachℍ, “refuge” etc.), and the UK (e.g., “United Kingdom”, “England”, “Wales”, “Scotland”, “London” etc.). The search terms will be combined using the Boolean operators ‘AND’ and ‘OR’. MeSH terms will also be included. The preliminary search strategy for MEDLINE can be found in Appendix 3.

Grey literature will also be searched for using a three-step strategy. Firstly, four specific grey literature databases will be searched: National Grey Literature Collection, ETHoS, Social Care Online and the Violence Against Women Network. Search terms will include: “domestic violence”, “sexual violence”, “service”, “support” and “intervention”. As above, only studies published after 01 January 1982 will be included. Secondly, a call for evidence will be circulated via email to a range of local and national services, service providers and research networks, to request any relevant service evaluations or reports that will be useful in answering the review questions and meet the inclusion criteria. Contacts will be emailed again if there is no initial response after two weeks. Finally, targeted website searching of relevant UK charities and organisations will take place. Websites will be searched for pages with links to reports, research, or publications. Titles and descriptions (where applicable) will be assessed according to the inclusion and exclusion criteria and potentially relevant documents will be downloaded. Where there are many pages of potentially relevant results, only the first five pages will be assessed. Where there is a search function, the following terms will be searched: “Service”, “Evaluation”, “Intervention” and “Report”. Backwards and forwards citation searching of all included studies will take place to supplement the database and grey literature searches to identify any further relevant studies.

## Screening of studies

Citations for all studies identified in the search will be transferred to Endnote to remove duplicates, before being uploaded into Rayyan software [44]. Study selection will be completed in two stages: first titles and abstracts will be screened according to the inclusion and exclusion criteria; next full texts will be screened to identify studies eligible for inclusion. Two reviewers will carry out screening independently. Any disagreement will be resolved by discussion, or with reference to a third reviewer if consensus cannot be reached. Reasons for exclusion of full texts will be recorded.

## Data extraction

Data extraction will take place using a piloted data extraction form and will focus on characteristics relevant to this review:

- Methods: study design, authors, date of publication, funding.
- Setting: country, sector.
- Participants: gender, age, ethnicity, immigration status, study inclusion and exclusion criteria, number of participants.
- Intervention: type of intervention or service, type of comparison, who it was delivered by.
- Outcomes: name of outcome, measure of effect (e.g., means and standard deviations, risk ratios, or number of events), time-points.

Two reviewers will be involved in data extraction. One reviewer will extract the data (SC), and a second will check the extracted data (AB). Any disagreements will be resolved through discussion. In cases where discussion does not resolve the disagreement, a third reviewer will be involved. Where data is missing, corresponding authors will be contacted and asked to supply said data.

## Quality appraisal

The risk of bias assessment tool will depend on the type of study. RCTs will be assessed using the Cochrane Risk of Bias 2 tool [45]. Non-randomised comparative studies and pre/post intervention studies will be assessed using the ROBINS-I tool [46]. Grey literature will be assessed using the AACDOS tool [47]. Assessments of bias will be done at the outcome level.

Two reviewers (SC and AB) will carry out risk of bias assessments independently. Assessments will be compared, and any disagreements will be resolved through discussion. In the case that discussion cannot resolve the disagreement, a third reviewer will be involved. The risk of bias ratings will not be used to inform the exclusion of studies. Rather, the assessments will be used to make a statement regarding the quality of the data.

## Data synthesis

A narrative synthesis will be conducted to summarise the included services and interventions, and their impact on the outcomes. The interventions and services will be described according to the template for intervention description and replication (TiDIER) [48] if / where appropriate, and grouped according to the different types of interventions (i.e., housing, advocacy, community-based, open-access, psychosocial support, legal support, financial support, MARACs, police-based services, and combinations).

Where the data permits it (i.e., there are ≥3 studies reporting the same outcome), meta-analyses will be conducted using Stata or Review Manager. A random effects model will be used to generate a pooled risk ratio or (standardised) mean difference. Heterogeneity will be assessed using the I2 statistic. Planned subgroup analyses will take place if heterogeneity is substantial or considerable (defined as I2 =50-90% and I2 =75-100%) [49]. This will include: study design; setting (third sector; private sector; public sector); relationship between the person who has experienced violence and the person who used violence (e.g., (ex)intimate partner; stranger; domestic but not partner; friend/acquaintance; professional; mixed/any); the population the service or intervention is aimed at (e.g., those who have experienced violence; perpetrators of violence; both); type of service or intervention provider (e.g., specialist DSVA; specialist but not DSVA; non-specialist); and type of violence (e.g., primarily DVA focused; primarily SVA focused; combined DSVA).

Additionally, sensitivity analyses will be conducted for the following: removal of high risk of bias studies, and removal of one study at a time. This will allow for exploration of potential biases. Funnel plots will additionally be produced to assess publication bias if more than 10 studies are identified.

In the case that meta-analysis is not appropriate, only the narrative synthesis will be presented and will follow the Synthesis Without Meta-analysis (SWiM) reporting guideline [50].

## Certainty in cumulative evidence

Where appropriate (i.e., where data have been pooled), a quality assessment using the GRADE framework will be used to assess the degree of certainty of the body of evidence. This takes into account risk of bias, inconsistency, imprecision, indirectness and publication bias.

## Discussion

This systematic review will provide an overview of the effect of UK support interventions and services for DSVA on the safety and wellbeing of those who have experienced DSVA. To our knowledge, this will be the first comprehensive review to collate and compare different types of support interventions and services in the UK context. By identifying the types of interventions and services producing the biggest impacts on these outcomes, we will help service providers and organisations across sectors to prioritise limited resources, as well as provide justification for securing additional funding for such activities.

The proposed systematic review has strengths that will enhance the comprehensiveness and robustness of findings. For instance, the inclusion of a comprehensive grey literature search strategy will allow for identification of reports and evaluations carried out by specialist support services that are not peer reviewed and may not be publicly available, thus reducing publication bias and allowing identification of maximal reports. This review also benefits from the continued contribution from specialist DSVA organisations who will provide guidance and advise on the development of the review, as well as aid in its dissemination. Finally, the review will be methodologically robust, following PRISMA guidelines and involving two reviewers throughout. However, there are also some limitations that will need to be considered when interpreting review findings. For instance, the nature of the literature is likely heterogenous in terms of both specific services and reporting of these services. This means that meta-analysis may be unlikely, limiting the review to a narrative synthesis. In such a case, narrative synthesis remains a useful and insightful approach, and thus we are confident that the proposed review will produce relevant and informative results. There are also several strengths and limitations of the evidence we expect to find. For instance, it is expected that much of the evidence will be based on service evaluations found in the grey literature. This means that much of the data will be based on real-world evidence, increasing external validity of review findings. However, as grey literature does not go through a peer review process, the quality of the evidence may be lower, therefore risk of bias assessments will be crucial.

## Patient and public involvement

An advisory group has been set up comprising six specialist DSVA organisations who are involved in the delivery, planning, funding or support of specialist support DSVA services in the UK. The group commented on and prioritised aspects to be the focus and scope of this review, including the population, types of violence and abuse, and defining of support services and interventions, as well as the design of this protocol. For example, as a result of the group’s feedback, the scope of the review was broadened from support services and interventions provided only by DSVA organisations, to include any support services and interventions for people who have experienced DSVA regardless of UK setting. They will continue to be involved throughout the review process to consult on data extraction, interpretation and dissemination of findings.

## Ethics and dissemination

Ethical approval is not applicable for this study since no original data will be collected. The results will be disseminated through peer-reviewed publications, and conference presentations, and non-academic outputs tailored to end user groups. Our advisory group will be consulted for additional sources through which to disseminate findings.

## Authors’ contributions

All authors contributed to the conceptualisation of the study protocol. SC produced the first draft of the study protocol and the manuscript. All co-authors reviewed the manuscript and approved the final version.

## Data Availability

No datasets were generated or analysed during the current study. All relevant data from this study will be made available upon study completion.

